# Feasibility and acceptability of smartphone technology for patients to self-record vital signs in the emergency department (The FacED Study): a study protocol

**DOI:** 10.1101/2025.05.25.25328325

**Authors:** Jared Charlton-Webb, Kathryn Willis, Gabriel Jones, Sarah White, Caspar Norris, Manesha Bageya, Heather Jarman

## Abstract

**Introduction:** Overcrowding and nursing workload are two major issues within urgent and emergency care (UEC) settings and contribute to incompletion or omission of patient vital signs in initial assessment and patient monitoring. This could potentially have a negative impact on patient health outcomes. Software enabling contactless monitoring of patient vital signs using camera photoplethysmography (PPG) has been developed, where patients can measure their own vital signs by recording a 30-second video on their smartphone on the software platform. This software could prove beneficial within UEC settings to help to reduce nursing workload and allow prioritisation of higher order tasks and could help to reduce long wait times for initial triage assessment. Before any such software can be implemented, it is first necessary to assess the feasibility and acceptability of the software in situ among patients in UEC settings, as well as the acceptability among staff working in triage.

**Methods and Analysis:** To assess feasibility of the software, 1500 patients attending three UEC services at St George’s University Hospitals NHS Foundation Trust will be invited to take part in a study where vital signs will be measured manually and using smartphone PPG software. Feasibility will be measured using a survey after data collection. Staff acceptability will be measured using a short survey among 20-40 staff. Patient acceptability of digital health technology in UEC settings will be measured by a questionnaire among 10,000 patients.

**Ethics and Dissemination:** This research has been ethically approved by the NHS London – Surrey Research Ethics Committee and the Human Research Authority (reference: 25/PR/0222; IRAS project ID: 346745). Informed consent will be obtained prior to the participant undergoing any activities that are specifically for the purposes of the study. Findings from this research will be disseminated via journals and conferences and used to inform further research in the field.

**Strengths and Limitations:**

- This observational study aims to assess the feasibility of using novel photoplethysmography (PPG) technology to measure vital signs in the Emergency Department
- Whilst several studies have explored the use of PPG, FacED will ascertain if this can be correctly used by patients in the ED
- The study includes several outcomes and will generate data on the acceptance of digital technologies in ED care
- The ability to recruit from three urgent and emergency care (UEC) settings within one NHS Trust is a strength of this research as it will enable a wide range of participants to be recruited from across emergency care, making the study target sample sizes more attainable and will provide feasibility assessments in 3 locations.
- This feasibility study will test the appropriateness of the methods to answering the study question but at present, the study materials are only accessible to English-understanding patients and data will be collected on participant’s first language

## Introduction

One of the biggest issues facing Urgent and Emergency Care Settings (UEC) internationally is crowding, defined as when “the demands on an ED exceed the capacity”.(1) Crowding is known to lead to long wait times, delays for assessment, increased patient morbidity and poor health outcomes.(2,3) Whilst several strategies to reduce delays to assessment have been proposed, including frontloading triage teams and FastTrack assessment,(1,4) the issue of ongoing monitoring and reassessment of patients in crowded waiting rooms has yet to be clearly tackled.

A key component of patient assessment and reassessment in UECs is the recording of vital signs. Patient vital signs include heart rate, blood pressure, oxygen saturation, temperature, and respiratory rate. In UEC vital signs are recorded at least once, firstly as part of an initial assessment for patients’ presenting with illness conditions,(5) and if needed, to monitor for clinical deterioration or improvements.(6) Despite the need to complete timely and regular recording of vital signs there is evidence that factors such as heavy nursing workload and ED crowding leads to incomplete or complete omission of recording.(7,8)

One potential solution that may ease nursing workload caused by high numbers of patients and low throughput is the implementation of patient self-recorded vital signs using smartphone camera-based photoplethysmography (PPG). This may have utility for both initial assessment and ongoing monitoring of vital signs in stable patients, reducing the duration of the triage assessment and allowing nursing staff to focus on higher order tasks. Previous literature has implied high acceptability among ED patients with regards to using mobile devices to access and enter their personal and healthcare information. (9,10)

Camera PPG software to measure vital signs has been explored in various healthcare settings including cardiology,(11) geriatrics,(12) paediatrics,(13) and intensive care.(14) Research within UEC settings reveals that camera PPG software could prove successful within ED triage,(15,16) however previous work has focused on piloting the accuracy of the application(12,15,16) using small sample sizes and has not been widely trialled in a real-life uncontrolled scenario with a large population.(17)

### Aims and Objectives

The main aim of the study is to determine the feasibility and acceptability of patient facial self-recorded vital signs for adult walk-in patients in the ED. The objectives are to

- Explore the performance of facial recording of vital signs compared to standard methods
- Determine clinician acceptability of facial recording of vital signs in the ED

A secondary objective is to establish whether patients in urgent and emergency care settings are be interested in, or willing to use, mobile health technologies in their care.

## Methods

### Study Design

This is a prospective, multi-site study comprising three inter-related work packages: feasibility and acceptability of self-recording of vital signs by patients (WP1), staff acceptability (WP2), and patient acceptability of mobile health technology in their care (WP3). The protocol was developed according to the Strengthening of Reporting of Observational Studies in Epidemiology (STROBE) statement.(18)

### Setting

The study will take place across three sites - the Emergency Department (ED), Urgent Treatment Centre (UTC) and Enhanced Primary Care Hub (EPCH) as part of St George’s University Hospitals NHS Foundation Trust. Recruitment will take place from 01 June 2025 – 31 December 2026. The study sites are situated in two different geographical locations in Southwest London.

### Participants

Participants will be identified by members of the clinical or research teams working in any of the sites. Screening for WP1and WP3 will use electronic medical records to establish absence of serious medical condition. Once identified potential participants will be approached by members of the research team and screened against the eligibility criteria (table 1), given information about the study and if in agreement complete written informed consent. For WP2 (clinical staff questionnaire) we will email eligible staff members to request participation.

**Table 1:** Eligibility criteria for the FacED feasibility study.

|  | WP1 + WP3 (patients) | WP2 (staff) |
| --- | --- | --- |
| Inclusion criteria | 'Walk-in' adult patients ≥ 18yrs in ED or UTC | Clinical staff (eg nurse, doctor) working in triage |
|  | • Has access to own smartphone<br>• Willing and able to give informed consent for participation in the study | or assessment area of either ED or UTC |
| Exclusion criteria | • Clinical condition warranting immediate medical attention | • Unwilling to complete questionnaire |

For WP3, participants will also be recruited through QR codes on posters in waiting rooms at participating sites. The QR code will direct a participant to a questionnaire landing page which will include a link to the participant information sheet. The participant will then click to confirm consent statements to provide informed consent to the survey. Patient-identifiable information is not collected during this survey as it is anonymous.

### Study size

WP1: the smartphone vital signs recording work package will use convenience sampling. The sample size was calculated to estimate the intraclass correlation coefficient (ICC) with a high level of precision. Assuming an expected ICC of 0.75, a sample of 1,178 participants is required to achieve a 95% confidence interval with a total width of 0.025. To allow for missing data and withdrawals, 1,500 participants will be recruited to ensure adequate power for assessing agreement between the smartphone-based and standard measures.

WP2: uses a non-probability convenience sample of clinical staff working in the assessment areas of each of the sites. We aim to obtain a representative clinical staff (by profession and experience) and will recruit between 20 and 40 members of staff.

WP3: uses a non-probability convenience sample of patients presenting to the ED over the recruitment period. There is no sample size calculation for the survey on digital health use in the ED patients. Reason for attendance is not a factor in inclusion and based on previous work using similar recruitment methods we anticipate that 10,000 participants will be recruited over the study duration.

### Variables

The outcome variables to assess the study aims are as follows:

- Feasibility: defined as the proportion of patients consented who were able to complete the vital signs recording using patientcheck.in
- Acceptability of self-recorded vital signs (patients): affinity for technology use - how comfortable, confident, and willing participants are when using digital or technological tools; and acceptability and patient experience of use of PPG application
- Performance: reliability of the platform to accurately measure the vital signs (blood pressure, heart rate, respiratory rate, oxygen saturation) recorded using a web-based PPG application compared to near simultaneous measurement of vital signs using a standard monitor
- Acceptability (clinical staff): electronic survey of personal technical affinity and a usability evaluation of the patient-recorded vital signs

### Data sources / measurement

The data collected in this study is anonymised and all participants will be assigned a unique identification (ID) number.

For WP1, participants will be asked to complete two types of data collection. A flow chart for WP1 can be found in **figure 1**.

**Fig 1:**
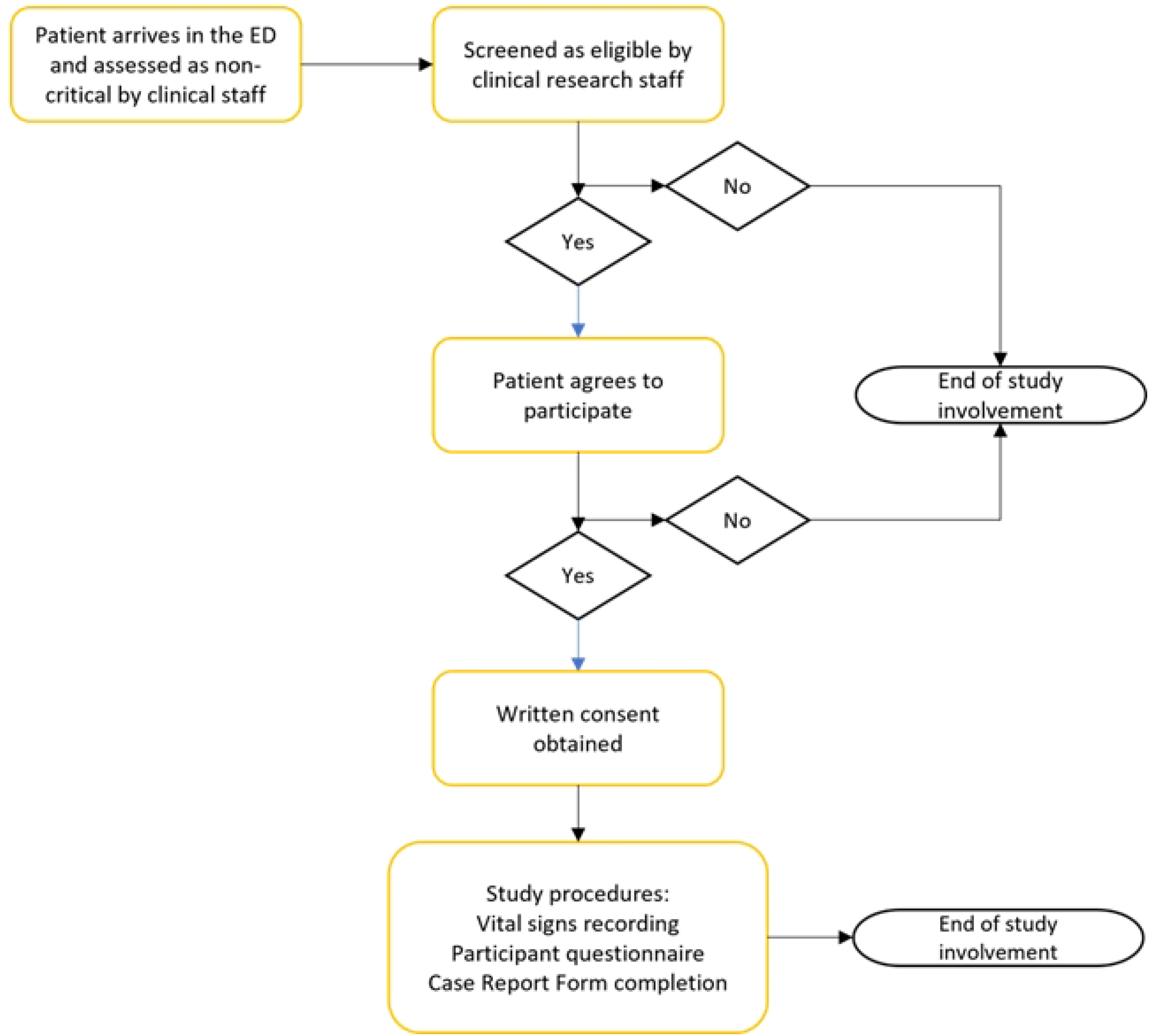
Study flow diagram depicting the recruitment process for WP1

- A clinical research nurse will support the patient through the data collection phases if necessary. Completion of their vital signs using PPG technology on their smartphone accessed through a QR code link on a webpage. The webpage will not capture any personal information, and all data is removed once the participant leaves the website. Participants will be guided through the capture of their vital signs by a research nurse if needed. Clinical or research staff will near-simultaneously complete the patients vital signs using traditional methods.
- Post-PPG recording participant acceptability data will be captured electronically using an online questionnaire. Participants will be able to complete the survey on their personal devices, following a linked QR code or can be provided with a handheld electronic device. The electronic survey contains the invitation to participate letter (participant information sheet), consent form and the questionnaire. Advantages of the electronic format include ease of administration, ease of data collection and reduced environmental impact versus the use of paper. The questionnaire has been designed to meet the study objectives, drawing from previous literature on digital technology acceptability and technical affinity. The questionnaire is comprised of 17 questions. There are 4 demographic questions, Fitzpatrick skin tone scale, and smartphone operating system. 8 questions including The Single Ease Question (SEQ) and System Usability Scale (SUS)(19,20) will be used to measure the usability of the software, and 5 questions will measure self-reported technical affinity (21). The survey should take no longer than 5 minutes for each participant to complete, and participants will be notified of this at the start of the survey.
- Participant deprivation index and ED length of stay information will be captured from the hospital electronic patient record (EPR) by the research team and linked to the patient acceptability data following consent.

To determine levels of clinical staff acceptability of the use of PPG technology (WP2), participants will complete an electronic questionnaire designed to capture workplace demographics (profession and seniority), 5 questions will measure self-reported technical affinity(21) and 10 questions to determine the perceived usefulness of the PPG technology on their practice.(22)

Finally, to establish whether patients in urgent and emergency care settings are be interested in, or willing to use, mobile health technologies in their care (WP3) we will collect data on age, gender, ethnicity, 1st language, educational attainment level and smartphone ownership. Self-reported technical affinity will be measured with 5 questions (21) and rating questions on areas where participants feel it is acceptable to use digital health technologies as part of their care. It will take participants approximately 15 minutes to complete.

### Bias

Bias will be minimised in the study by collecting data through standardised reporting and assessment of measures and using a recognised evaluation questionnaires tool. All research staff collecting data will be provided with study specific training on procedures and data collection.

### Analysis and statistical methods

The electronic surveys will be hosted on the REDCap Platform under a password-protected personal account held only by the Chief Investigator and study team. REDCap provides compliance with General Data Protection Regulations and is ISO 27001 compliant. A link to the REDCap Data protection policy is provided within the participant information sheet.

Descriptive statistics will be used to characterise participants by age, gender, highest educational attainment, IMD and ethnicity. Feasibility will be reported with a percentage and 95% CI. Acceptability to patients as measured by SEQ and overall SUS score will be summarised, using mean and standard deviation (SD). The association between acceptability to patients and age, educational attainment, IMD and ethnicity will be assessed using regression methods. Descriptive statistics will be used to summarise the findings of the acceptability to clinical staff survey.

Performance of the app will be assessed using the intraclass correlation coefficient (ICC) for the whole sample and for a priori defined subgroups. ICC’s take values between 0 and 1 where, 0 implies no agreement and 1 is perfect agreement and represents the proportion of the total variability in the observations that is due to the differences between pairs(23). Bootstrapped confidence intervals will be calculated for estimates of ICC for each subgroup to ensure the robustness of the analysis on smaller samples.

### Patient and Public Involvement (PPI)

Effective patient and public involvement is of key importance to providing safe and effective emergency care. Discussions with ED service users informed the study design. Feedback emphasized the importance of clear instructions for internet application use and the brevity of questionnaires.

### Ethics and dissemination

The study obtained UK NHS ethical approval by the London – Surrey Research Ethics Committee (REC number 25/PR/0222) on 4^th^ April 2025. Informed consent will be obtained prior to participants undergoing any activities that are specifically for the purposes of the study. Consent for WP1 will be taken electronically by a member of the research team. For WP2 and WP3 consent is ‘self-completed’ online and requires no input from the clinical or research teams. Research staff will not be aware of who has taken part and will not be able to identify any participants. There is no follow-up or other direct contact by research staff.

Findings from this research will be shared and discussed with the study steering committee in preparation of wider dissemination through journals and conferences.

## Data Availability

No datasets were generated or analysed during the current study. All relevant data from this study will be made available upon study completion.

## Author Contributions

HJ conceived the study design and wrote the study protocol. JCW and KW wrote the first draft of this paper, refined by HJ. All authors are co-applicants on the grant submission and reviewed the manuscript. HJ is responsible for data integrity and will act as a guarantor.

## References

1. Royal College of Emergency Medicine. The Management of Emergency Department Crowding. 2024 [cited 2025 Feb 28]; Available from: https://rcem.ac.uk/wp-content/uploads/2024/01/RCEM_Crowding_Guidance_Jan_2024_final.pdf

2. Mackway-Jones K, Marsden J, Windle J. Emergency Triage: Manchester Triage Group [Internet]. 3rd ed. 2013 [cited 2025 Feb 28]. Available from: https://books.google.co.uk/books?id=DZusAQAAQBAJ&dq=K.+Mackway-Jones,+J.+Marsden+and+J.+Windle,+Emergency+Triage:+Manchester+Triage+Group,+3rd+ed.,+BMJ+Books,+2023.+&lr=&source=gbs_navlinks_s

3. Royal College of Physicians. National Early Warning Score (NEWS) 2: Standardising the assessment of acute-illness severity in the NHS [Internet]. 2017 [cited 2025 Feb 28]. Available from: https://www.rcplondon.ac.uk

4. Redfern OC, Griffiths P, Maruotti A, Recio Saucedo A, Smith GB. The association between nurse staffing levels and the timeliness of vital signs monitoring: A retrospective observational study in the UK. BMJ Open. 2019;9(9).

5. van der Linden MC, Meester BEAM, van der Linden N. Emergency department crowding affects triage processes. Int Emerg Nurs. 2016 Nov 1;29:27–31.

6. Selvaraju V, Spicher N, Wang J, Ganapathy N, Warnecke JM, Leonhardt S, et al. Continuous Monitoring of Vital Signs Using Cameras: A Systematic Review. Sensors [Internet]. 2022 Jun 1 [cited 2025 Feb 28];22(11):4097. Available from: https://www.mdpi.com/1424-8220/22/11/4097/htm

7. Bautista M, Cave D, Downey C, Bentham JR, Jayne D. Clinical applications of contactless photoplethysmography for vital signs monitoring in pediatrics: A systematic review and meta-analysis. J Clin Transl Sci [Internet]. 2023 May 25 [cited 2025 Mar 3];7(1):e144. Available from: https://pmc.ncbi.nlm.nih.gov/articles/PMC10310860/

8. Yu X, Laurentius T, Bollheimer C, Leonhardt S, Antink CH. Noncontact Monitoring of Heart Rate and Heart Rate Variability in Geriatric Patients Using Photoplethysmography Imaging. IEEE J Biomed Health Inform. 2021 May 1;25(5):1781–92.

9. Capraro GA, Balmaekers B, den Brinker AC, Rocque M, DePina Y, Schiavo MW, et al. Contactless Vital Signs Acquisition Using Video Photoplethysmography, Motion Analysis and Passive Infrared Thermography Devices During Emergency Department Walk-In Triage in Pandemic Conditions. J Emerg Med. 2022 Jul 1;63(1):115–29.

10. Kobayashi L, Chuck CC, Kim CK, Luchette KR, Oster A, Merck DL, et al. Comparison of Video Photoplethysmography, Video Motion Analysis, and Passive Infrared Thermography against Traditional Contact Methods for Acquiring Vital Signs in Emergency Department Populations. 2023 IEEE 14th Annual Ubiquitous Computing, Electronics and Mobile Communication Conference, UEMCON 2023. 2023;134–41.

11. Mather JD, Hayes LD, Mair JL, Sculthorpe NF. Validity of resting heart rate derived from contact-based smartphone photoplethysmography compared with electrocardiography: a scoping review and checklist for optimal acquisition and reporting. Front Digit Health. 2024 Feb 29;6:1326511.

12. UK Mobile Phone Statistics 2024 -Stats Report - Uswitch [Internet]. [cited 2025 Feb 28]. Available from: https://www.uswitch.com/mobiles/studies/mobile-statistics/

13. Kim E, Torous J, Horng S, Grossestreuer A V., Rodriguez J, Lee T, et al. Mobile device ownership among emergency department patients. Int J Med Inform. 2019 Jun 1;126:114–7.

14. Goldfine CE, Knapp A, Goodman GR, Hasdianda MA, Huang H, Marshall AD, et al. Media and technology usage and attitudes in emergency department patients. Front Digit Health. 2022 Oct 31;4:894683.

15. Brooke J. SUS: A ‘Quick and Dirty’ Usability Scale. In: Usability Evaluation In Industry [Internet]. CRC Press; 1996 [cited 2025 Feb 28]. p. 189–95. Available from: https://www.taylorfrancis.com/chapters/edit/10.1201/9781498710411-35/sus-quick-dirty-usability-scale-john-brooke

16. Doll WJ, Xia W, Torkzadeh G. A confirmatory factor analysis of the end-user computing satisfaction instrument. Doll WJ, Xia W, Torkzadeh G, editors. MIS Q. 1994;18(4):461.

17. Fritsch SJ, Blankenheim A, Wahl A, Hetfeld P, Maassen O, Deffge S, et al. Attitudes and perception of artificial intelligence in healthcare: A cross-sectional survey among patients. Digit Health [Internet]. 2022 Aug 8 [cited 2025 Feb 28];8. Available from: https://journals.sagepub.com/doi/10.1177/20552076221116772

18. Bonett DG. Sample size requirements for estimating intraclass correlations with desired precision. Stat Med [Internet]. 2002 May 15 [cited 2025 Feb 28];21(9):1331– 5. Available from: https://onlinelibrary.wiley.com/doi/full/10.1002/sim.1108

